# High-dimensional deconstruction of pancreatic ductal adenocarcinoma identifies tumor microenvironmental communities associated with survival

**DOI:** 10.1101/2022.04.29.22274376

**Authors:** Erik P. Storrs, Abul Usmani, Prathamesh Chati, Ian Sloan, Bradley A. Krasnick, Ramandeep Babbra, Peter K. Harris, Faridi Qaium, Deyali Chatterjee, Chris Wetzel, S. Peter Goedegebuure, Thomas Hollander, Hephzibah Anthony, Jennifer Ponce, Shahed Badiyan, Lauren Henke, Hyun Kim, David G. Denardo, Gabriel D. Lang, Natalie D. Cosgrove, Vladimir M. Kushnir, Dayna S. Early, William G. Hawkins, Ashiq Masood, Li Ding, Ryan C. Fields, Koushik K. Das, Aadel A. Chaudhuri

## Abstract

Bulk and single-cell analyses of the pancreatic ductal adenocarcinoma (PDAC) tumor microenvironment (TME) have revealed a largely immunosuppressive milieu. Thus far, efforts to utilize insights from TME features to facilitate more effective therapeutics have largely failed. Here, we performed single-cell RNA sequencing (scRNA-seq) on a cohort of treatment-naive PDAC time-of-diagnosis endoscopic ultrasound-guided fine needle biopsy samples (n=22) and surgical samples (n=6), integrated with 3 public datasets (n=49), resulting in ∼140,000 individual cells from 77 patients. Based on expression markers assessed by Seurat v3 and differentiation status assessed by CytoTRACE, we divided the resulting tumor cellular clusters into 5 molecular subtypes based on expression of previously reported marker genes: Basal, Mixed Basal/Classical, Classical Low, Classical High, and ADEX. We then queried these 5 tumor cell profiles, along with 15 scRNA-seq-derived tumor microenvironmental cellular profiles, in 391 bulk expression samples from 4 published datasets of localized PDAC with associated clinical metadata using CIBERSORTx. Through unsupervised clustering analysis of these 20 cell state fractions representing tumor, leukocyte and stromal cells, we identified 7 unique clustering patterns representing combinations of tumor cellular and microenvironmental cell states present in PDAC tumors. We termed these cell state patterns communities, and found them to correlate with overall survival, tumor ecotypes, and tumor cellular differentiation status. The community associated with worst overall survival contained basal tumor cells, exhausted CD4 and CD8 T cells, and was enriched for fibroblasts. In contrast, the highest overall survival was associated with a community high in immune cell enrichment. The differentiation state of tumor cells (assessed by CytoTRACE) was also correlated with survival in a dose-dependent fashion. Further, we identified a subset of PDAC samples that were significantly enriched for CD8 T and plasma cells that achieved a 2-year overall survival rate of 71%, suggesting we can identify PDAC patients with significantly improved prognoses and, potentially, higher sensitivity to immunotherapy.

In summary, we identified novel tumor microenvironmental communities from high-dimensional analysis of PDAC RNA sequencing data that reveal new connections between tumor microenvironmental composition and patient survival that could lead to better upfront risk stratification and more personalized clinical decision-making.

## INTRODUCTION

Pancreatic ductal adenocarcinoma (PDAC) is the third leading cause of cancer death in the United states with a 5-year survival rate of 10.8%^1^. PDAC has remained largely refractory to available therapeutics, with a hallmark of heterogenous chemotherapeutic responses in subsets of patients^2^. Over the past decade, bulk tumor sequencing has enabled annotation of the genomic landscape in pancreatic ductal adenocarcinoma (PDAC)^3,4^. This has led to several classification systems for PDAC^2,4,5^. The general consensus consistently demonstrates the existence of two major subtypes of PDAC: the classical or pancreatic progenitor subtype associated with a relatively better prognosis (characterized by differentiated ductal markers like PDX1) and the basal-like, squamous or quasi-mesenchymal subtype associated with a poorer prognosis (characterized by the expression of basal-like markers like cytokeratin 81 (KRT81))^3,4^. While these insights have allowed for the elucidation of unique transcriptional networks^6,7^, they have yet to allow for the development of effective clinical interventions.

Underlying this, in part, is the fact that these subtyping techniques rely on gross analysis of bulk sequencing data, creating blind spots in individual cell states and features of individual cells within a single tumor sample. This issue is especially pronounced in PDAC, where only 20% of cells are tumor cells, and thus the ability to fully decipher all cellular variants is limited when using traditional next generation sequencing (NGS) methodologies^3^. Underscoring the importance of granular analysis, recent experiments have demonstrated the complex interplay of subtypes of PDAC tumor cells with the associated stroma and the inflammasome^8^. Advances in single cell RNA sequencing (scRNA-seq) have provided the added ability to describe individual cell profiles and query individual cell states^9–12^. These insights enable a more in-depth analysis of the tumor microenvironment (TME) and tumor heterogeneity with unparalleled granularity. Indeed, several scRNA-seq efforts have demonstrated that PDAC tumors are a heterogeneous and spatially diverse admixture of “basal-like” and “classical” cells with potential for plasticity between transcriptomic states with unknown prognostic implications^13–15^. The majority of these efforts have focused on the utilization of surgical resection specimens^9,13^ or biopsies of patients with liver metastases^14^ or potentially from archival material^16^, rather than focusing on time-of-diagnosis endoscopic ultrasound (EUS) needle biopsies in resectable or locally advanced patients. Integrating these technologies with standard of care tissue acquisition at the time of diagnosis may allow these granular insights to serve as potential biomarkers for personalized therapeutics.

Here, we perform scRNA-sequencing of PDAC from 28 standard endoscopic ultrasound guided fine needle biopsy (EUS-FNB) specimens at the time of diagnosis and from surgical samples from tumor resections. We then integrate these cases with 49 samples from four additional publicly available bulk expression studies. Utilizing this approach, we identify TME cell states and previously described major molecular subtypes^3^ in our single cell data. We then apply the CytoTRACE algorithm, an innovative method for inferring developmental cell states, including potential cancer stem cells, which reveals a developmental dichotomy within classical tumor cells^17^. To clinically translate the cell states found in our single cell expression data to patient outcomes, we use CIBERSORTx, a method for granularly inferring cell state abundances, in 391 PDAC patients with tumor bulk RNA-seq and microarray samples, to infer 20 cell state fractions in four publicly available datasets^3–5,18^. We then used unsupervised clustering techniques and EcoTyper^19^ to identify “communities” and ecotypes that we find to be strongly associated with overall survival. Our innovative approach reveals new predictive insights into PDAC starting from the time of diagnosis, thus paving the way toward personalized risk-adapted therapy based on upfront granular genomic cell state analysis.

## METHODS

### PDAC tumor processing

Endoscopic ultrasound was performed on patients with suspected solid pancreatic masses based on CT or MRI imaging (**Figure 1, Supplementary Table 1)**. The diagnosis of pancreatic adenocarcinoma was confirmed by formal pathologic evaluation. After clinical diagnostic tissue acquisition was completed with 2-3 passes of a 22-gauge needle, an additional pass was obtained with a backfin “fine-needle biopsy” (FNB) needle. Tissue was carefully washed with cold PBS, collected in RPMI 1640 media (Gibco) on ice when processed fresh or collected in freezing media (90%FBS + 10% DMSO, when processed at later time point) and dissociated into single cell suspension both mechanically and enzymatically as previously described^20^. Resected surgical tumor tissue was also dissociated in a similar way to obtain single cell suspension. Subsequently, single-cell suspensions were diluted to a final concentration of ∼1,000 cells/μl and sequencing libraries prepared using the 10x Genomics Chromium Single Cell 5’ library platform. Complementary DNA libraries were then sequenced on an Illumina NovaSeq S4 flow cell with a target of 50,000 reads/cell.

**Figure 1:**
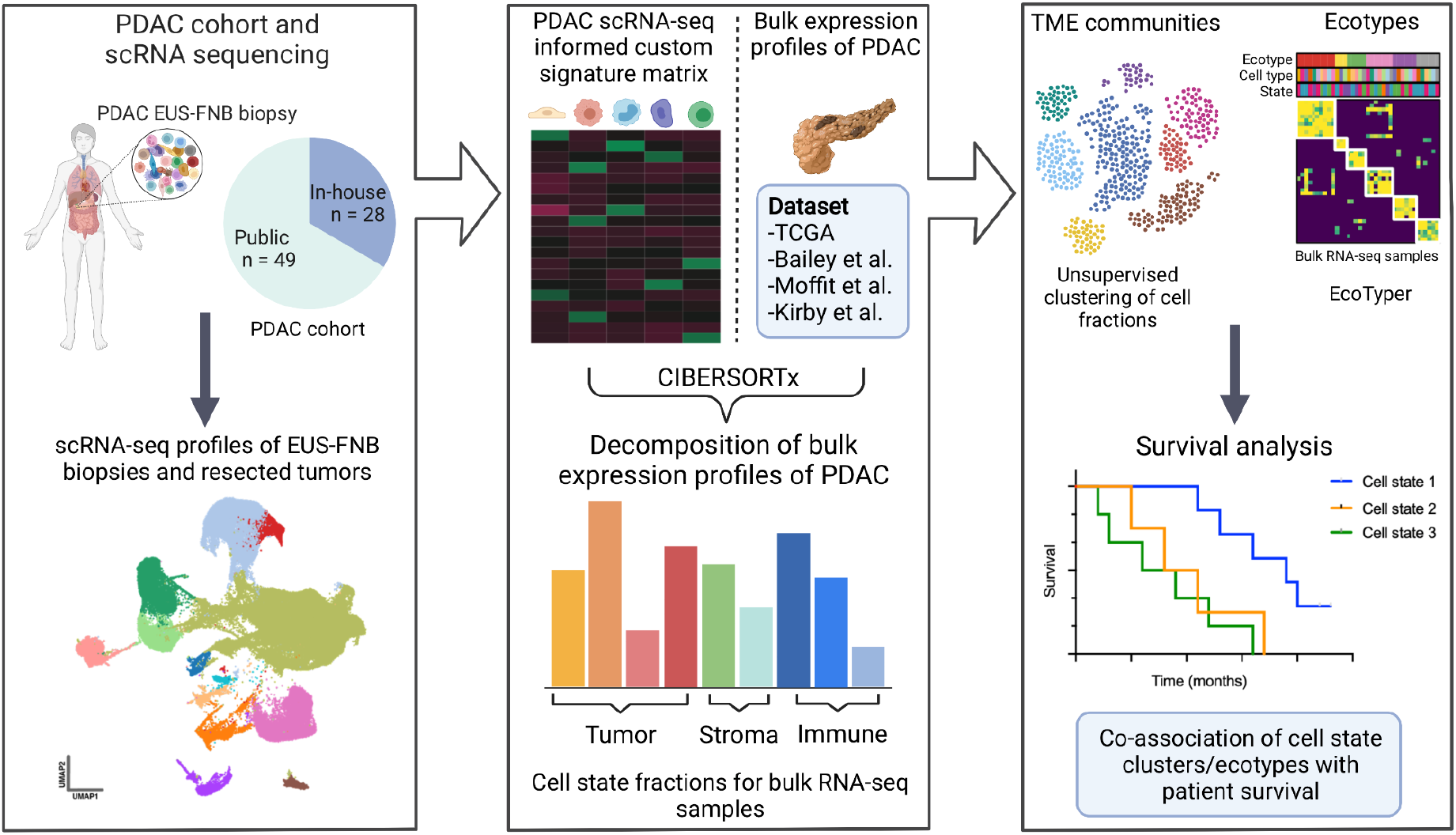
Study schema. Single-cell RNA-seq (scRNA-seq) was performed on treatment-naïve pancreatic adenocarcinoma (PDAC) tumor samples acquired by EUS-FNB (n=22) or surgical resection (n=6). These were integrated with samples from three publicly available PDAC scRNA-seq datasets resulting in a combined dataset of ∼150k cells from 77 independent patient samples. The resulting data were used to identify PDAC cell states, including malignant and immune subtypes based on gene sets and known expression markers from published studies. With single-cell annotations in hand, we used the digital deconvolution tool CIBERSORTx to infer cell state fractions across 391 independent PDAC tumor samples across four publicly available bulk expression datasets. We then utilized unsupervised clustering, and orthogonally applied EcoTyper, to identify patterns of cell states in PDAC with prognostic significance.

### In-house scRNA-seq data processing

We aligned reads to the GRCh38 reference genome and obtained gene expression counts using 10x Cell Ranger 3.0.2 with default parameters^21^. FASTQ files were aligned to the GRCh38 reference genome with the STAR aligner^22^. Cell-specific unique molecular identifiers (UMIs) were then used to generate gene expression matrices. Cells that expressed less than 200 total genes and genes that were expressed in fewer than 3 cells were filtered from the dataset. Additionally, cells with a feature count of over 7,500 or a mitochondrial DNA percentage of over 20% were also filtered from the dataset.

### Integration of public scRNA-seq datasets

Filtered in-house EUS-FNB and surgical samples (n=28) were integrated with three publicly available scRNA-seq datasets. These datasets include Peng et al. (n=24)^10^, Lin et al. (n=10)^12^, and Chan-Seng-Yue et al. (n=15)^11^. Peng et al. was downloaded from the Genome Sequence Archive under project PRJCA001063, Lin et al. was downloaded from the GEO database at accession number GSE154778, and Chan-Seng-Yue et al. from the EGA under accession code EGAS00001002543 (**Supplementary Table 2**). Expression counts were integrated using anchor transfer methodology implemented by the Seurat single cell library^23^. First we normalized each dataset with the SCTransform function. We then identified the 3,000 most variable features to use for anchor transfer with the function SelectIntegrationFeatures and performed dataset integration using the functions FindIntegrationAnchors and IntegrateData.

### Cell state identification

The first 30 principle components (determined using RunPCA) of the integrated dataset were used for nearest neighbor computation, UMAP dimensionality reduction, and clustering. The Louvain algorithm was used to cluster single cell expression data via the FindClusters Seurat function. For initial clustering a resolution of .75 was used. Clusters were merged and assigned to cell types based on known cell type expression markers: CAF (BGN+, FAP+), NK/T-cell (CD45+, CD3G+ and/or NKG7+), DC (FCER1A+, CD74+, HLA-DRA+), Malignant (EPCAM+, KRT18+), Endothelial cell (PECAM1+), Erythrocyte (HBA1+), B cell (MS4A1+, KIT+), Mast cell (CPA3+), Monocyte (FCER1A+, CD14+, LYZ+), Plasma cell (SDC1+), Acinar (PRSS1+, CDH5+), Stellate (RGS5+). The NK/T cell cluster was further clustered with a resolution of 0.75. The resulting clusters were grouped into CD4 T cells (CD3G+, CD4+), CD8 T cells (CD3G+, CD8A+), CD4/CD8 T cells - exhausted (CD3G+, LAG3+, PDCD1+), NK (NKG7+, GNLY+), and T-reg (FOXP3+).

### Identification of PDAC malignant subtypes

Malignant cells were clustered with a resolution of 0.1. They were then labeled based on subtype markers previously described in the literature. We used 3 gene marker sets (Bailey et al., Moffitt et al., and Collisson et al.)^2,4,5^ for cluster assignment. For each subtype in Bailey and Moffitt we used the top 20 published genes. From the Collisson gene set we used all available genes. The genes used for scoring each subtype are available in the supplemental materials (**Supplementary Table 8**). We then scored cells based on mean expression for genes in the marker gene sets for each subtype in the studies listed above. Malignant cell clusters in our single cell data were then assigned labels (Basal, Basal/Classical, Classical Low, Classical High, ADEX) based on enrichment for these marker gene sets.

### Bulk Expression Data Acquisition

TCGA PDAC clinical and bulk RNA-seq expression data were downloaded from the NCI GDC(https://portal.gdc.cancer.gov/); we restricted ourselves to samples used in Raphael et al. (n=136)^3^. Bailey et al.^5^ (n=87) were downloaded from the ICGC Data Portal (https://dcc.icgc.org/projects/PACA-AU). Moffit et al.^4^ (n=123) microarray expression and clinical data were downloaded from the Gene Expression Omnibus under the accession number GSE71729. Kirby et. al.^18^ was downloaded from the GEO databank under GSE79670 (**Supplementary Table 3**).

### Bulk expression deconvolution

Signature matrices were derived from our annotated single cell expression data, which were input as raw counts as previously described in Newman et al^24,25^. Bulk RNA-seq data, prior to CIBERSORTx^24^ input, were normalized to the total number of counts (CPM) for RNA-seq data (TCGA, Bailey, and Kirby), while microarray data (Moffit) was kept as downloaded.

CIBERSORTx was run in containerized form via Docker with the following parameters: --single_cell TRUE --rmbatchSmode TRUE.

### Tumor microenvironmental community analysis

Deconvolved cell fractions for the publically available bulk datasets demonstrated batch effects. In order to integrate samples from these datasets, we thus applied a clustering workflow inspired by scRNA-seq based batch correction and clustering methods. Specifically, after imputing cell fractions with CIBERSORTx, we applied batch correction to the samples from the different datasets to account for technical differences between them. To this end, we integrated the cell fractions by projecting them to principal component space with components fit with cell fractions from samples in Bailey et al. For plotting applications, the resulting principle components were then decomposed to 2D space with the UMAP algorithm (https://arxiv.org/abs/1802.03426). Following dimensionality reduction, samples from Bailey et al. were then clustered with the Leiden algorithm^26^ to assign initial communities. The clusters identified by the Leiden algorithm were then transferred to other datasets via the k-nearest neighbors (KNN) algorithm. This functionality was implemented in Scanpy^27^ with the following functions and parameters: scanpy.pp.neighbors(n_neighbors=10), scanpy.pp.pca, scanpy.tl.umap(spread=5.), scanpy.tl.leiden(resolution=.8), and scanpy.tl.ingest functions. Clusters were then given community labels based on their enriched cell states: Basal, Mixed, Classical Low, Classical High, ADEX, Mixed - Immune High, and Mixed - Stromal High

Enrichment for cell fraction was computed via the Wilcoxon rank-sum test. Significance levels were adjusted via the Benjamini-Hochberg method, and implemented with the scanpy.tl.rank_genes_groups function.

### Ecotype discovery

Ecotype discovery was performed with EcoTyper^19^. Cell states and ecotypes were discovered in the TCGA bulk RNA-seq dataset according to steps in EcoTyper documentation Tutorial #4: De novo Discovery of Cell States and Ecotypes in Bulk Expression Data (https://github.com/digitalcytometry/ecotyper). Briefly, cell fraction estimation was performed with CIBERSORTx using scRNA-seq expression profiles from cell states identified in the *Cell state identification* methods section. To allow EcoTyper to discover its own malignant cell states, we labeled all malignant cells as malignant, rather than their subtype-specific classification. Following the discovery of cell states and ecotypes on TCGA bulk expression samples, steps for EcoTyper cell state and ecotype recovery were performed by following the steps in Tutorial #1: Recovery of Cell States and Ecotypes in User-Provided Bulk Data. These recovery steps were executed for Bailey, Moffit, and Kirby datasets.

Overlap of bulk samples in discovered ecotypes and communities was computed as the ratio of the number of samples present in both ecotype and community / total number of samples in the ecotype.

### Survival analysis

All Kaplan-Meier curves were generated with the Python lifelines package. For survival analysis of Malignant cell fractions and EcoTyper-discovered cell states, log rank p-values were computed with the lifelines logrank_test function.

### Gene set enrichment analysis (GSEA)

Pathway enrichment analysis for genes significantly associated with the S01 CD8 T cell state was done with ToppFun^28^. Significant GO: Molecular Function^29^ pathways were selected based on enrichment of the top 20 cell state associated genes. Top pathways were then rank-ordered by their -log10 FDR corrected p-values.

### Development state analysis

CytoTRACE (v0.3.3) was used to determine developmental states of malignant cells in the scRNA-seq expression dataset^17^. The scRNA-seq expression matrix was normalized to CPM and run with Scanorama^30^ batch correction according to steps listed in the Custom Integrated CytoTRACE tutorial (https://cytotrace.stanford.edu/).

Malignant subtype developmental status was determined in bulk RNA-seq data by correlating the top 20 genes for each EcoTyper-discovered malignant cell state with our malignant subtype single cell data.

## RESULTS

### Quantification of malignant and tumor microenvironment cell states in integrated scRNA-seq datasets

We performed scRNA-seq of PDAC from standard-of-care endoscopic ultrasound guided fine needle biopsy (EUS-FNB) specimens at the time-of-diagnosis and from surgical samples obtained from tumor resections to enable a clinically integrated, comprehensive view of PDAC (**Figure 1, Supplementary Table 1**). In total, we acquired 28k cells across 22 independent PDAC samples for our in-house EUS-FNB cohort, and 20k cells from 6 independent PDAC samples for our in-house surgical cohort. To increase power, we then combined the in-house scRNA-seq data with three publicly available datasets: Peng et al, Lin et al, and Chan-Seng-Yue et al.^10–12^ (**Supplementary Table 2**). Following integration of our single cell data with Seurat^23^, we increased our sample size to a total of 141k cells from 77 independent PDAC tumors. The integrated dataset was clustered and annotated based on known cell type markers, where a total of 13 cell types were found (**Figure 2.A**). With the exception of erythrocytes, every cluster had representation from multiple datasets, indicating that dataset-specific batch effects were largely removed during integration (**Supplementary Figure. 1.A**).

**Figure 2:**
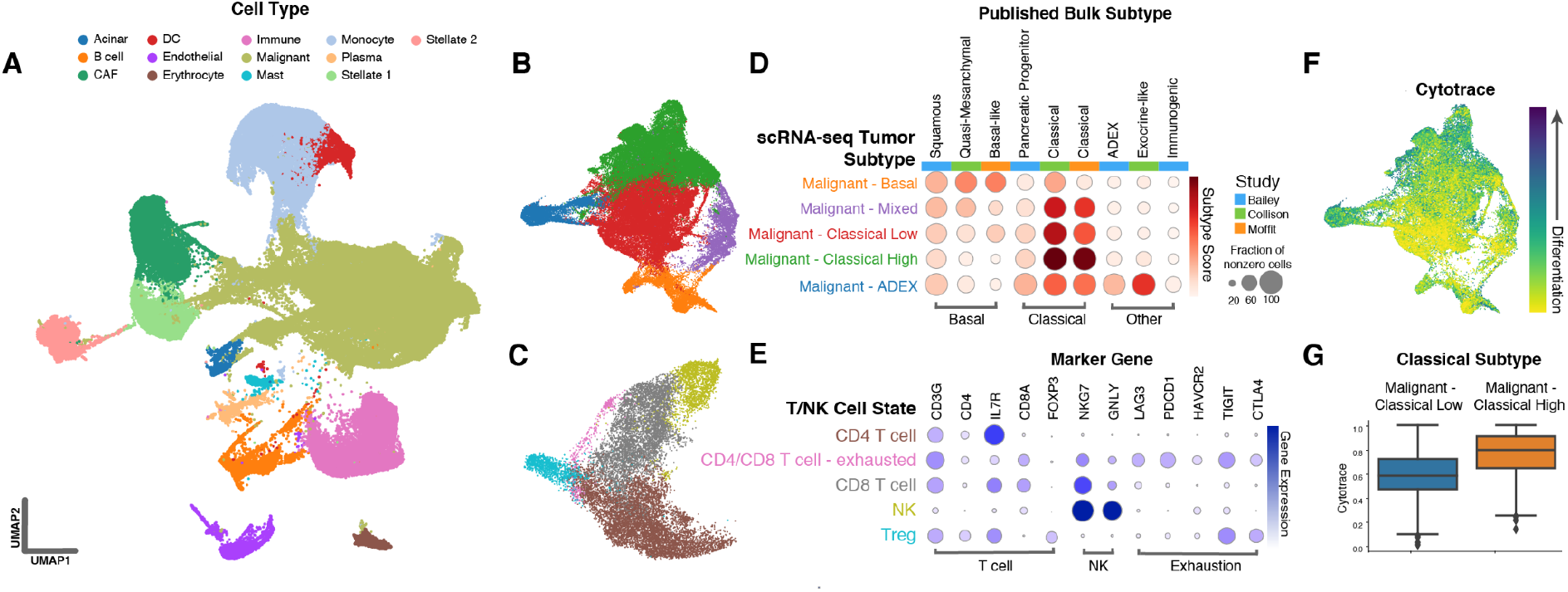
Single-cell dissection of pancreatic ductal adenocarcinoma. **A)** UMAP decomposition of scRNA-seq expression profiles from ∼150k cells representing 14 cell types (from 77 independent PDAC tumors). UMAP decomposition of **B)** 5 malignant subtypes and **C)** 5 immune cell states. **D)** Comparison of our scRNA-seq-derived malignant subtype expression profiles to published bulk expression subtypes. **E)** Comparison of scRNA-seq immune cell states to known expression markers. **F)** CytoTRACE differentiation continuum across the malignant single-cell subtypes. **G)** CytoTRACE developmental scores differ between the two subtypes of Classical tumor cells identified in our scRNA-seq data.

The Malignant and T cell clusters were further subclustered into more fine-grained cell states. For the malignant cluster, we identified five total subclusters (**Figure 2.B**). These subclusters were categorized based on aggregate expression of markers from three previous studies: Bailey et al., Collison et al., and Moffit et al. (**Figure 2.D, Supplementary Table 8)**. When the expression signatures derived based on these known subtypes were overlaid on the malignant cell subclusters, we found remarkably good concordance with the known subtypes (**Supplementary Figure. 2.A-D**). Using gene expression microarray data, Moffitt et. al. categorized PDAC into classical and basal-like populations^4^. Bailey and Collision later used bulk RNA sequencing data to describe squamous, endocrine, pancreatic progenitor and immunogenic (Bailey), and quasi-mesenchymal, classical and ADEX (Collison) subtypes of PDAC^2,5^. The consensus subtypes found in these studies are thus 1) Classical (Classical, pancreatic progenitor), and 2) Basal (basal, squamous-like, quasi-mesenchymal).

Some studies have called the existence of the immunogenic and ADEX/endocrine subtypes into question^3^, however, in addition to Basal and Classical subtypes, we also observed a cluster overexpressing genes associated with the ADEX/endocrine subtype in our single cell data. Based on strong concordance with the known subtypes, we annotated the five malignant subclusters as the following: Basal, Mixed, Classical Low, Classical High, and ADEX. In addition to further clustering the malignant cells, we also further subdivided the T lymphocyte cluster into five states: CD4 T cell, CD8 T cell, exhausted T cell, NK, and Treg (**Figure 2.C, E**). For all the above cell states we observed good representation from multiple datasets, indicating none of the clusters are an artifact of a particular dataset. (**Supplementary Figure. 1. B,**).

We then sought to determine the developmental status of the malignant subclusters. For this, we used CytoTRACE^17^ to obtain developmental scores for each tumor cell, with cells having a high CytoTRACE score being less developmentally mature, and those with a low score being more differentiated. We found two of the five malignant subclusters, ADEX and Classical High, were more differentiated than the other subclusters (**Figure 2.F, G**). This developmental continuum between Classical and Basal clusters has been previously shown, as the Basal subtype is known to more likely undergo the epithelial-mesenchymal transition (EMT), resulting in higher rates of metastasis^31,32^. However, it is interesting that the “less classical” of the two classical subclusters is more poorly differentiated than the other, perhaps indicating that this subcluster carries more stem-like features, and as a result is more aggressive than its more developmentally mature counterpart.

### Community identification in clinical bulk expression datasets

We then extended our granular single cell expression profiles to publicly available bulk expression datasets with associated clinical metadata (**Supplementary Table 9**). To this end, we applied the digital deconvolution tool CIBERSORTx to obtain cell state proportions for each bulk sample (**Figure 3.A**). First, we derived a signature matrix from expression profiles of each annotated cell state in the single cell RNA-seq dataset. Then, using CIBERSORTx, we deconvolved 391 samples from four separate bulk expression datasets: Moffit et al., Bailey et al., Kirby et al., and TCGA^3–5,18^ (**Supplementary Table 7**). After cell state fractions were imputed, we sought to identify clusters of samples with similar cell state fraction profiles. We term these clusters of samples “communities”. After batch-correcting sample-level cell fractions to account for differences in data type and methodology between the bulk datasets, we clustered the samples with the graph-based Leiden^26^ clustering algorithm. Following clustering, we were left with seven distinct communities, each with representation from at least two of the bulk expression datasets (**Figure 3.B**).

**Figure 3:**
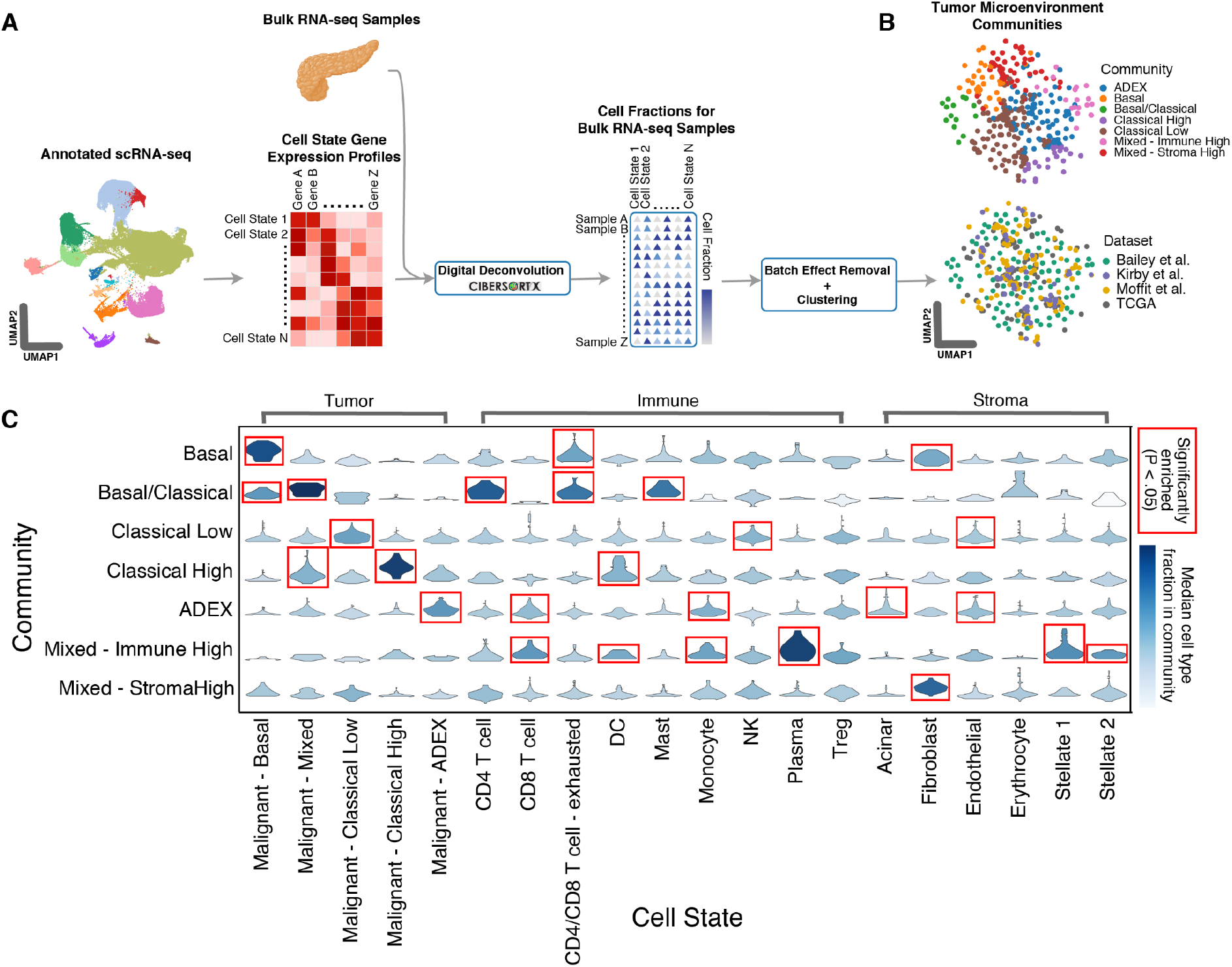
Unsupervised discovery of tumor microenvironmental communities within PDAC bulk expression data. **A)** Schema for generating the expression signature matrix from scRNA-seq-derived cellular subtypes followed by deconvolution of bulk expression data with CIBERSORTx. **B)** UMAP decomposition of deconvolved bulk expression samples colored by tumor microenvironmental community and dataset. **C)** Plot showing cell state fractions in each TME community. Red boxes represent significantly enriched malignant, immune, or stromal cell fractions (*P* < .05).

The communities were then labeled based on patterns of cell fraction enrichment (**Figure 3.C, Supplementary Table 4**). Specifically, there were four communities that fell on a spectrum between the main basal and classical PDAC subtypes. These communities were termed Basal, Basal/Classical, Classical Low, and Classical High. We also found a community enriched with the ADEX malignant subtype cell fraction, as well as two communities with higher proportions of stromal/immune cells, which we name Mixed - Stromal High and Mixed - Immune High. Interestingly, we also observed an overrepresentation of exhausted T cells in the Basal and Basal/Classical communities (**Figure 3.C**).

We then orthogonally validated the discovered communities with the in-silico TME dissection tool EcoTyper^19^. Similar to our unsupervised community detection approach, EcoTyper takes as input single cell expression profiles. These profiles are then used to impute gene expression and identify cell states for samples in bulk RNA-seq and microarray datasets (**Figure. 4.A**). Additionally, EcoTyper groups significantly associated cell states into Ecotypes, which are an analogous abstraction to our TME communities. When EcoTyper was applied to the 391 samples from the bulk expression datasets, we found six distinct ecotypes, labeled E1-E6, each with its own distinct pattern of cell state enrichment (**Figure 4.B**). Ecotype E1 is highly enriched for immune cell states and has low tumor purity. Further, it displays high levels of enrichment for Bailey et al. immunogenic samples^5^, which are samples known to have high ratios of immune cells^3^. Further indicating an immune-enriched makeup, ecotype E1 exhibited highly specific overlap with the Mixed - Immune High community (**Figure 4.C**). In addition to ecotype E1, ecotypes E3 and E6 showed high overlap with the Basal and Basal/Classical communities, respectively, while ecotypes E2, E4 and E5 did not show specific association with any of the identified communities.

**Figure 4:**
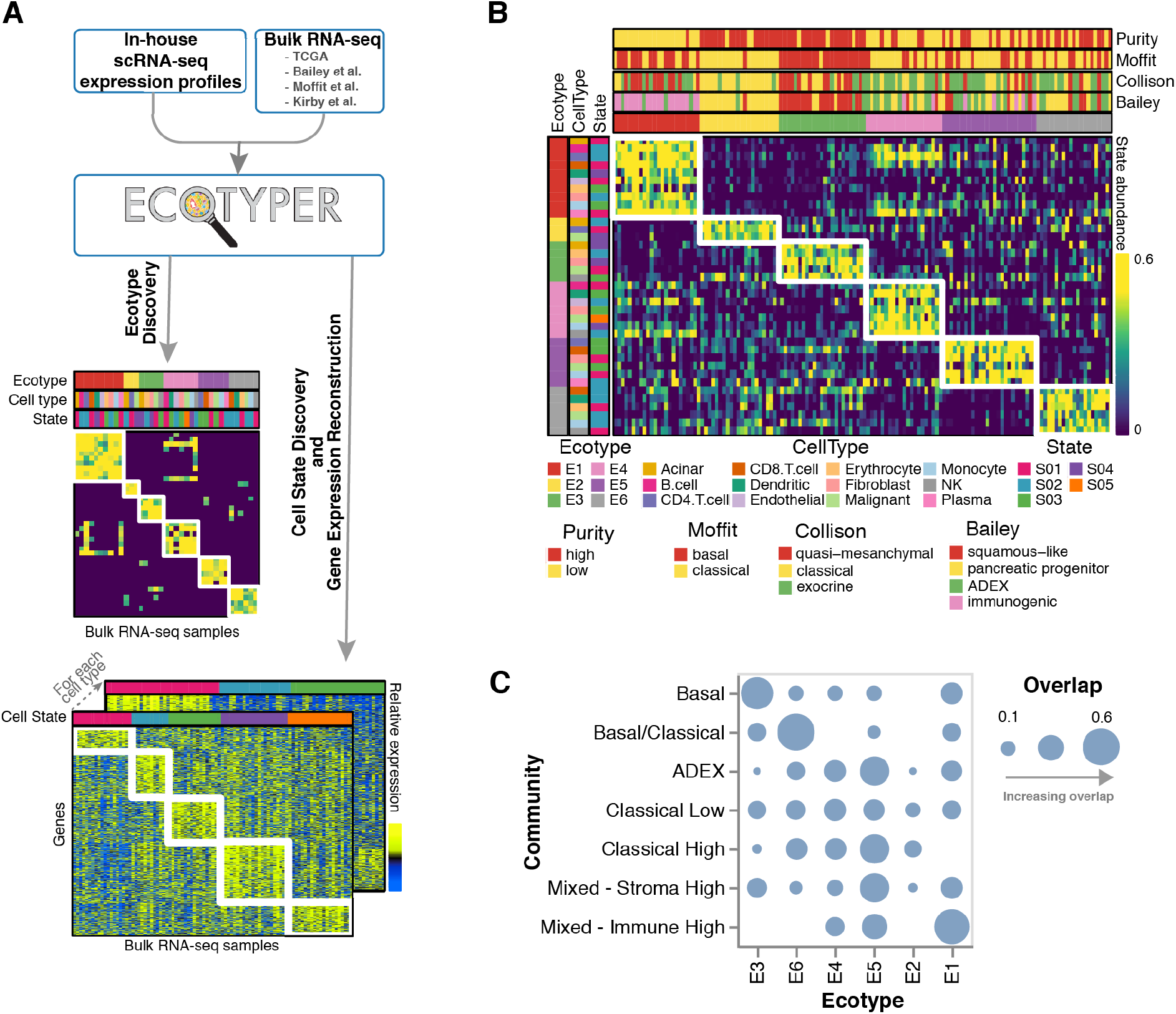
Discovery of PDAC ecotypes with Ecotyper. **A)** Expression profiles from in-house scRNA-seq data were used to discover cell state fractions and reconstructed gene expression profiles for samples in four publicly available bulk expression datasets. **B)** Heatmap showing ecotype cell state abundances for each bulk expression sample across six discovered ecotypes. **C)** Overlap between ecotypes and TME communities identified within bulk expression samples.

### Patterns of TME cell state association are indicative of patient outcome

We then performed survival analysis on the cell states that make up the above communities and ecotypes. As expected, we found there to be a spectrum of survival related to the abundance of malignant subtypes, with the most basal cell states showing the poorest survival, while the most classical subtypes displayed the greatest overall survival (**Figure 5.A**). Meanwhile, intermediary/mixed subtypes fell between the two. This relationship between basal and classical subtypes has been described extensively in the literature^14^. This trend appeared in both the deconvolved cell state fractions comprising the TME communities, as well as EcoTyper-discovered malignant cell state abundances. Of particular note, we also observed that the Classical Low cell state has worse survival than the Classical High cell state, indicating that the Classical subtype partitions into distinct survival subgroups, consistent with findings by Chan-Seng-Yue et al.^11^. Additionally, we found that EcoTyper-discovered CD8 T cell states are associated with survival, where the S01 CD8 T cell state is indicative of improved overall survival, while its S02 and S03 counterparts portend worse outcomes (**Figure 5.B**).

**Figure 5:**
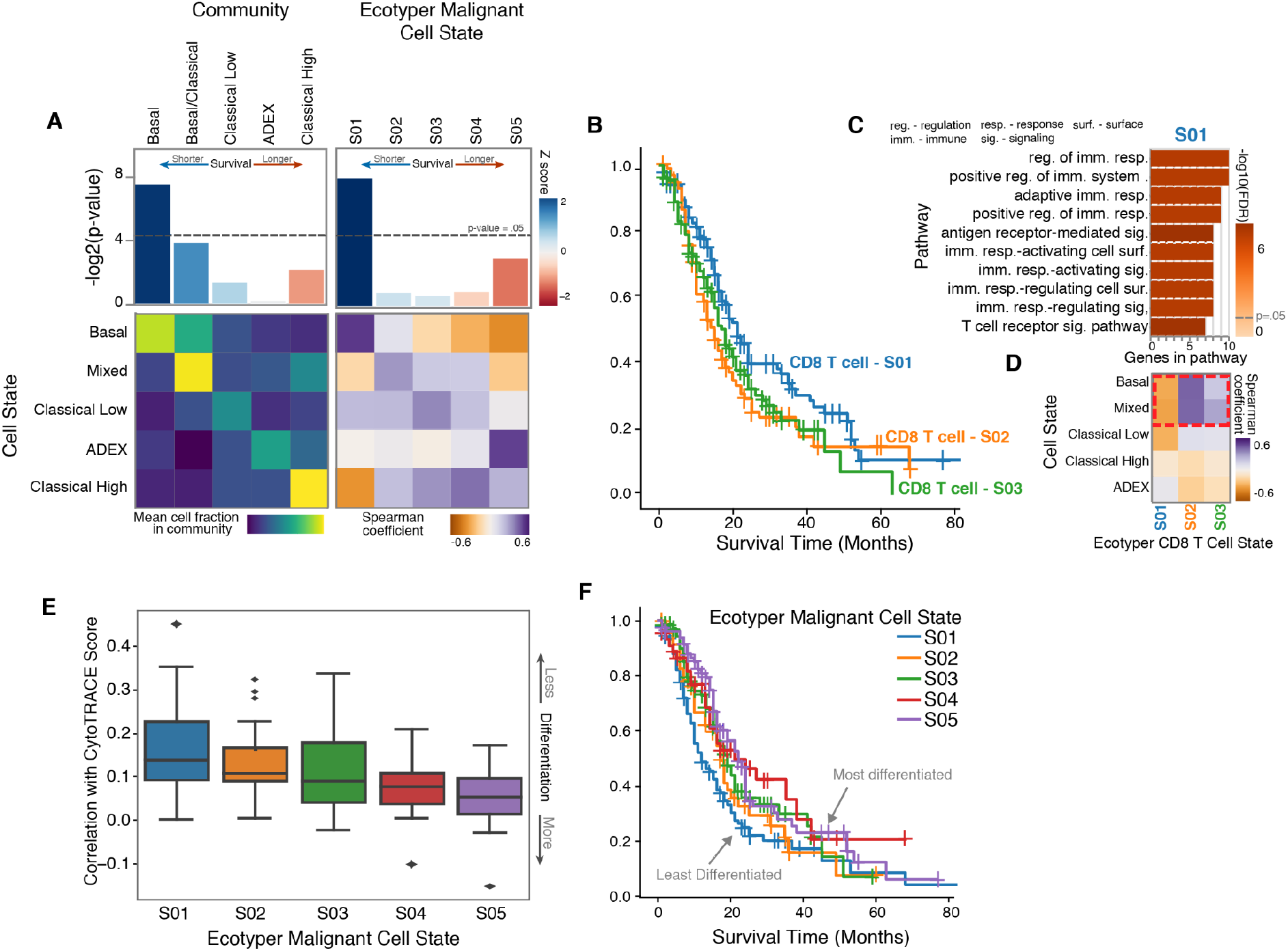
TME communities and ecotypes are associated with survival. **A)** Deconvolved cell state fractional abundances for each TME community in bulk expression data, correlated with EcoTyper-discovered malignant cell states. -log2 p-values for associations with overall survival are also shown. **B)** Kaplan-Meier curves of EcoTyper-discovered CD8 T cell states. **C)** Gene set enrichment analysis of EcoTyper-discovered CD8 T cell state with the highest overall survival shows enrichment for immune cell activating pathways. **D)** Correlation between deconvolved bulk malignant subtype cell fractions and EcoTyper-derived CD8 T cell states shows enrichment of S02/S03 in basal and mixed communities and absence of the immune-activating S01 cell state. **E)** Correlation of top 20 genes associated with each EcoTyper-discovered malignant cell state with differentiation level inferred by CytoTRACE. **F)** Kaplan-Meier curves of EcoTyper-discovered malignant cell states.

To identify pathways upregulated in S01, we performed a gene set enrichment analysis (GSEA) on the top 20 genes significantly associated with the S01 CD8 T cell state. We found that S01 is enriched for genes associated with T cell activation, suggesting that tumors with more activated CD8 T cells have better outcomes (**Figure. 5.C, Supplementary Tables 5 and 6**). Next, we correlated the abundance of the EcoTyper-discovered CD8 T cell states with the deconvolved malignant cell state fractions to see if the activated S01 CD8 T cell state was strongly associated with a particular malignant subtype. We found that the immune-active S01 CD8 T cell state is negatively correlated with the Basal and Basal/Classical cell states, while its counterparts S02 and S03 are positively associated with the basal subtypes (**Figure. 5.D**). These findings suggest that the more aggressive, EMT-like Basal and Basal/Classical communities have fewer activated CD8 T cells present in their TME compared to other PDAC malignant subtypes.

Next, we sought to compare the developmental status of each EcoTyper-discovered malignant cell state. To do so, we took CytoTRACE score correlations from our malignant single cell data for the top 20 genes associated with each malignant cell state and displayed their distribution across each EcoTyper malignant cell state. We found that there not only exists a survival continuum between malignant cell states, but also across the developmental continuum, with Basal being the least differentiated and Classical High the most differentiated (**Figure. 5.E**). This finding aligns well with the developmental analysis of our single cell data, where we found developmental differences from cells across different subtypes. We also performed survival analysis where we partitioned each sample into its most abundant EcoTyper malignant cell state, and strikingly found that the developmental continuum associated strongly with patient outcomes, with the least differentiated malignant cell state having the worst overall survival (**Figure. 5.F**).

We further queried survival at the community and ecotype levels, where we found that immune dominance correlated with higher overall survival. Specifically, TME patterns with enrichment of immune cells, including non-exhausted CD8 T cells, monocytes and plasma cells, such as the ecotype E1 and the Mixed - Immune High community demonstrated much improved survival over tumor cell, fibroblast and exhausted T cell dominant communities and ecotypes (**Figure. 6.A, B**). Additionally, the Mixed - Stromal High community that was enriched for various stromal states also showed modestly increased survival compared to more tumor-centric communities and ecotypes (**Figure. 6.C**). These findings align with previous studies showing that increased immune cell infiltration, particularly of CD8 T cells and plasma cells, portends better outcomes in a variety of cancer types^33–35^.

**Figure 6:**
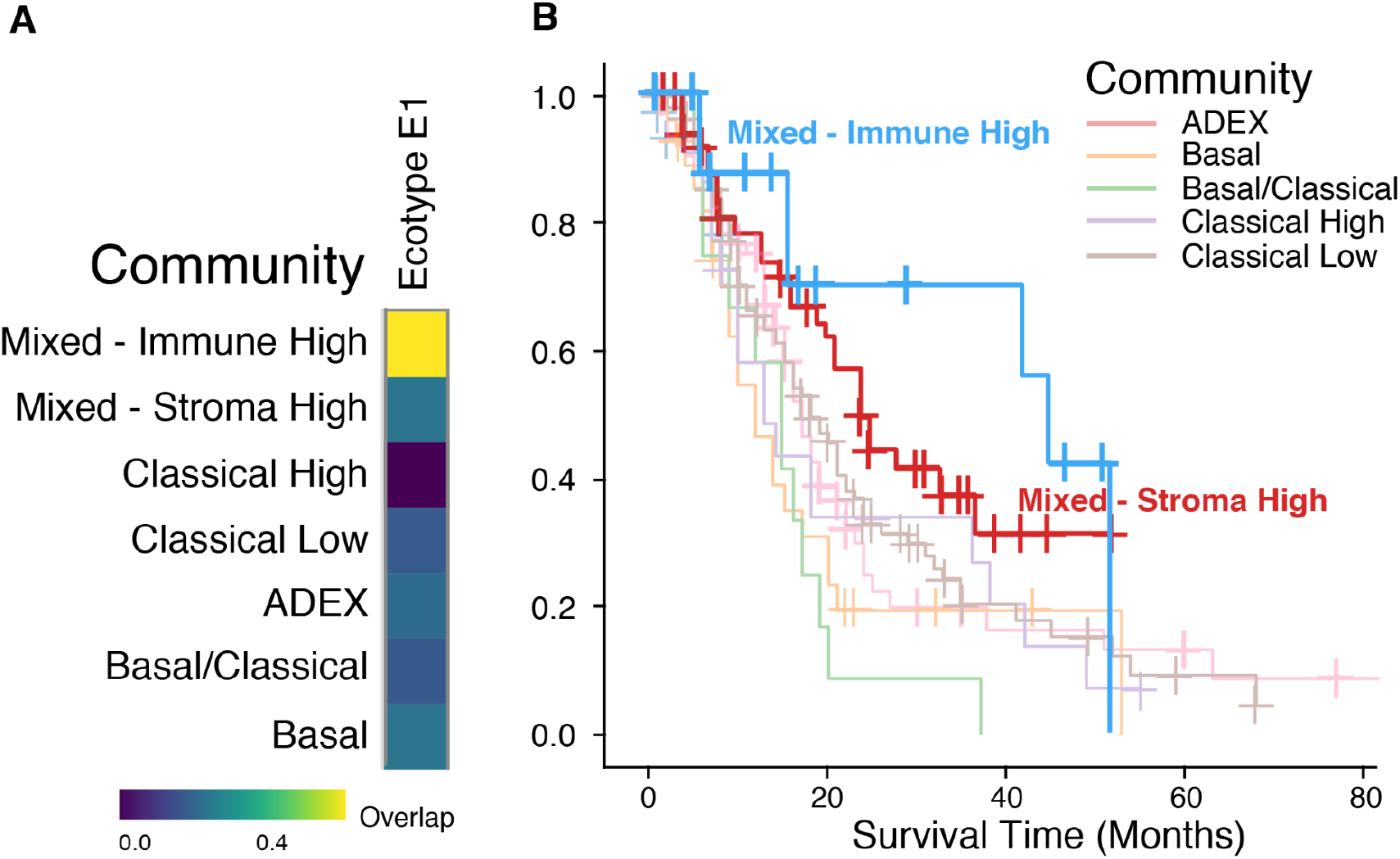
Higher overall survival in PDAC tumors with immune and stromally enriched microenvironmental communities. **A)** Overlap of Ecotype E1 with TME communities. Kaplan-Meier curves of **B)** TME communities showing increased overall survival in patients harboring immune- and stromal-high PDAC tumors.

## DISCUSSION

Using microarrays, Moffitt et. al. categorized PDAC into classical and basal-like populations^4^. Multiple groups have since performed bulk RNA sequencing to corroborate these findings and identify potential other tumor cell subtypes. Unlike bulk RNA-seq, scRNA-seq allows us to individually profile each cell, and thus appreciate the full breadth of diversity in cell types and profiles within the tumor microenvironment. This is especially important in a cancer like PDAC, where only ∼20% of cells in a biopsy sample are tumor cells, with the remaining cells representing various components of the TME^36^. Still, scRNA-seq is not practical yet within the clinical workflow. Thus, in this study, we followed our scRNA-seq analysis of 77 patients, including 28 obtained at the time of diagnosis, with bulk expression of 391 predominantly early-stage and surgically resected PDAC tumors followed by digital deconvolution.

By performing both scRNA-seq and bulk RNA-seq on such a large scale, we not only recapitulated known tumor subtypes (classical, squamous-like, and ADEX), but also uncovered evidence of tumor heterogeneity with several PDAC tumors harboring mixtures of subtypes (i.e., Mixed Basal/Classical). Furthermore, we discovered low and high classical malignant cell states, which correlated with a developmental dichotomy that we identified using CytoTRACE^17^. Using digital tissue deconvolution with CIBERSORTx, we then inferred 20 tumor and tumor microenvironmental cell state fractions across PDAC tumors from nearly 400 patients. With unsupervised clustering of these 20 different tumor/microenvironmental cell states, we identified cell state communities that were associated with overall survival. We then orthogonally validated these communities with the TME discovery tool EcoTyper^19^. Consistent with prior data, the basal community conferred a significantly worse prognosis as compared to the most classical community. Interestingly, other communities fell on a continuum between these two populations, and their ordering was based on factors such as TME makeup and dominant malignant subtype.

Additionally, survival correlated very strongly with malignant cell developmental status, suggesting that the communities associated with poorer survival are also more EMT-like. A recent article from Gulati et. al. demonstrated the ability to identify distinct developmental states within the luminal progenitor compartment of breast cancer using CytoTRACE, where knocking down genes associated with the immature malignant cell state led to decreased tumor growth *in vivo*^*17*^. As we also observed a striking association with survival in PDAC patients using CytoTRACE, similar methods could be applied to PDAC by targeting molecular pathways specific to immature malignant cell states, which could significantly improve clinical outcomes for otherwise high-risk patients.

We also found that 5% of pancreatic cancer patients harbor immune-enriched tumors with increased plasma cell, non-exhausted CD8 T cell and monocyte immune infiltration. These patients achieved strikingly higher overall survival than all other patients, with a 2-year overall survival of 71%. Indeed, tumor-infiltrating CD8 T and plasma cells have been shown to predict better survival in a variety of different solid tumor malignancies, especially in concert with immune checkpoint blockade^33,34^. It will be important to prospectively test whether these PDAC patients harboring immune-enriched tumors can be precisely selected using bulk RNA sequencing followed by digital deconvolution, and if their survival can be further enhanced with immune checkpoint blockade.

This study has several limitations. First, it utilized publicly available sequencing and clinical-correlative data. While all public data utilized for this work were previously published and also secondarily analyzed^2–5,10,12,18^, it will be important to corroborate our findings in a prospective setting. Second, scRNA-seq from time-of-diagnosis EUS-FNB samples is technically challenging given the limited sample material obtained. It will be important to corroborate our scRNA-seq findings by performing bulk RNA-seq (which can be performed with a lower sample amount) of EUS-FNB samples followed by digital deconvolution. Third, while we showed prognostic correlations between tumor microenvironmental features and overall survival, it remains unclear if these data have predictive potential. It will be important to perform clinical trials in the future where, for example, patients with immune-enriched tumors are selected to receive immune checkpoint blockade, or those with genomically less differentiated tumors are offered drugs targeting molecular pathways specific to immature malignant cell states.

In summary, we performed a large-scale transcriptomic and clinical-correlative analysis of 468 PDAC patients, including several fine-needle biopsy samples collected at the time of initial cancer diagnosis. Our high-resolution insights into the tumor microenvironment using scRNA-seq, bulk RNA-seq, digital deconvolution, community/ecotype analysis, and developmental state analysis have the potential to better personalize care and significantly improve outcomes for select patients.

## Data Availability

All data produced in the present study are available upon reasonable request to the authors

## Acknowledgements

We are grateful to the patients and families involved in this study, and to the clinical research team for collection of samples and clinical data. We also thank A. Newman for providing critical feedback on the manuscript. This work was supported by the National Institutes of Health (NIH) National Cancer Institute (NCI) Washington University SPORE in Pancreatic Cancer under award number 5P50CA196510 (W.G.H. and R.C.F.) including a Career Enhancement Program subaward to A.A.C. This work was also supported by the Washington University Human Tumor Atlas Research Center funded by the NCI under award number U2CCA233303 (L.D. and R.C.F.). This work was further supported by the NCI Washington University Participant Engagement and Cancer Genomic Sequencing Center under award number 1U2CCA252981 (L.D., R.C.F., A.A.C.), a NCI K08 career development award under award number K08CA238711 (A.A.C.), the Cancer Research Foundation Young Investigator Award (A.A.C.), the Washington University Alvin J. Siteman Cancer Research Fund (A.A.C.), and the V Foundation for Cancer Research V Scholar Award (A.A.C.). The funders played no role in study design, data collection and analysis, decision to publish, or preparation of the manuscript. This study also utilized the computational resources of the McDonnell Genome Institute at Washington University. Images from Biorender.com were used to create Figure 1.

## Author Contributions

E.S., A.U., P. C., R.C.F., K.D. and A.A.C. conceived of the study, developed strategies for related experiments, and wrote the paper. Data analysis and interpretation were performed by E.S., A.U. and P.C. with assistance from B.A.K., R.B. and P.K.H. Patient specimens were collected by A.U., I.S., B.A.K., P.K.H., C.W., S.P.G., T.H., H.A., G.D.L., N.D.C., V.M.K., and D.S.E., and were processed for expression profiling by A.U. Clinical characteristics and outcomes were determined by I.S., B.A.K., R.B., and K.K.D. Clinical data were curated by A.U., I.S., and K.K.D. All authors commented on the manuscript at all stages. K.K.D. and A.A.C. are co-senior authors and led all aspects of the project. All authors commented on the manuscript at all stages.

## SUPPLEMENTARY FIGURES

**Supplementary Figure 1:**
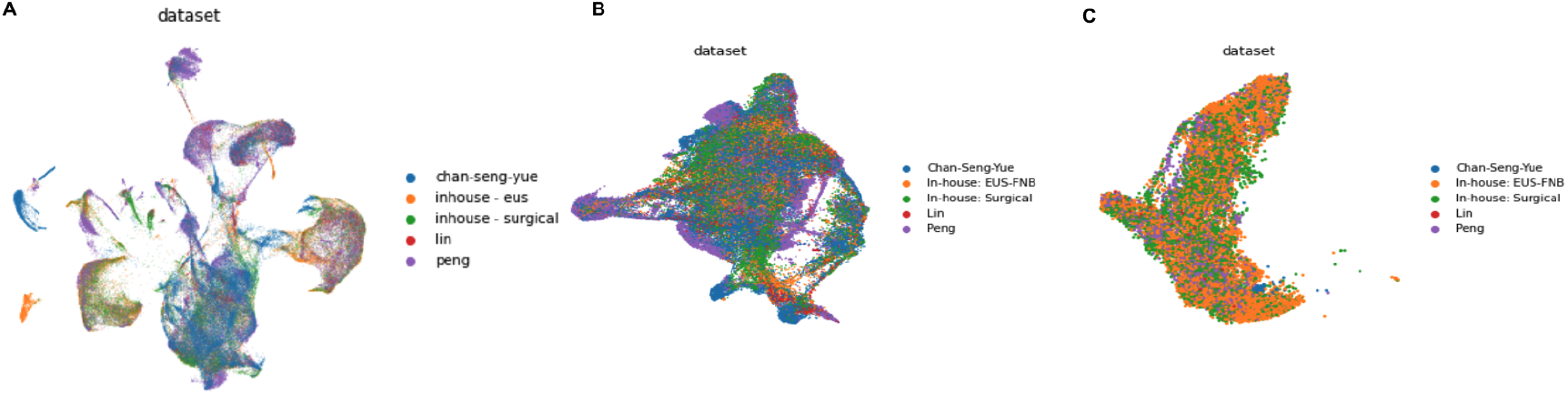
Dataset distribution in single cell data. UMAP-decomposed expression profiles overlaid with dataset membership for A) all cell types, B) malignant cells, and C) immune cells.

**Supplementary Figure 2:**
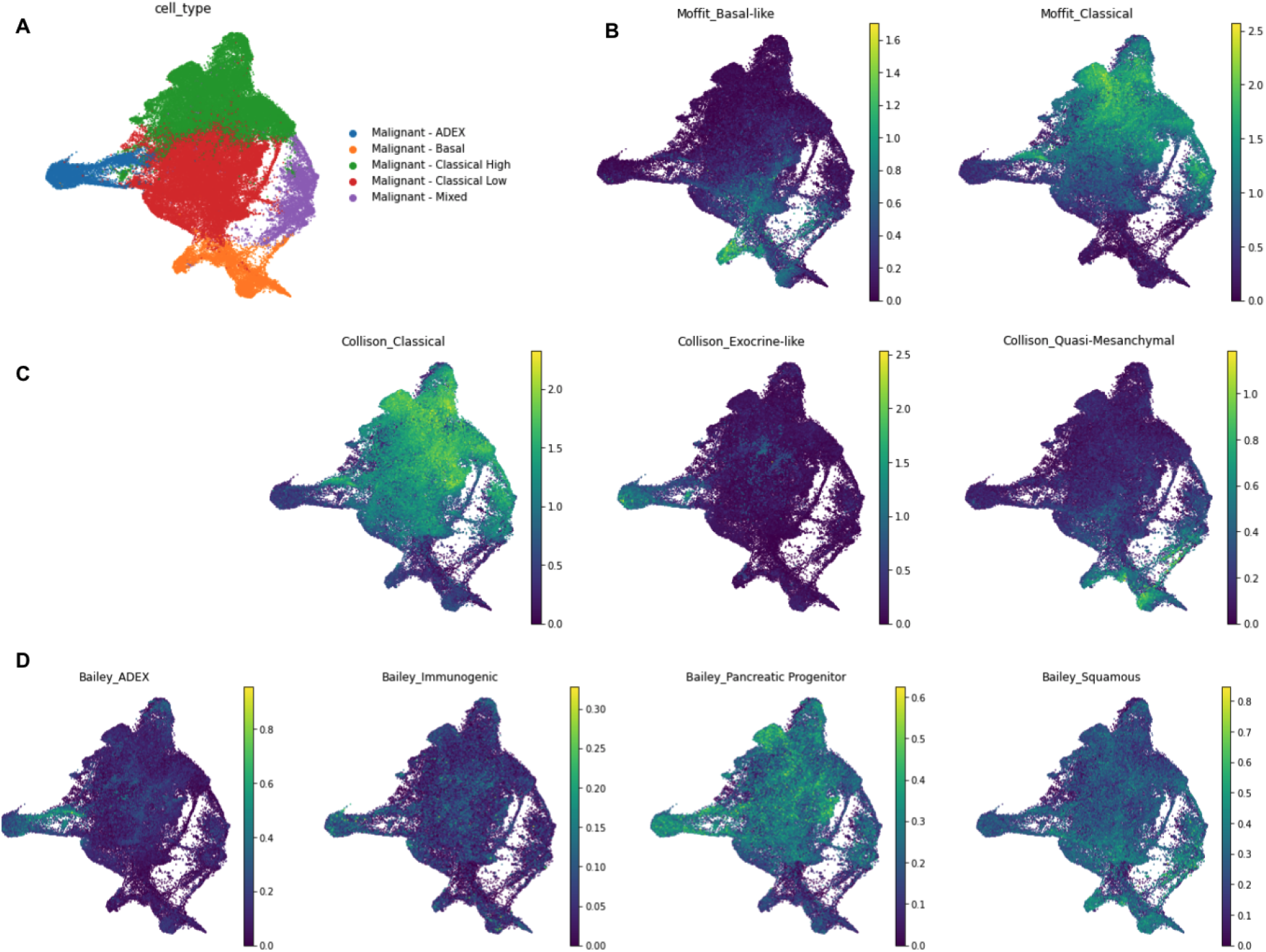
scRNA-seq subtype scores. UMAP-decomposed expression profiles overlaid with A) subtype classification, B) Moffit subtype scores, C) Collison subtype scores, and D) Bailey subtype scores.

